# HIV Dolutegravir Resistance and Multiclass Failure in Mozambique: Findings from a Real-World Cohort

**DOI:** 10.1101/2025.05.19.25327901

**Authors:** Fausto Ciccacci, Anna Maria Doro Altan, Noorjehan Majid, Stefano Orlando, Elton Umuasse, Marcia Rafael, Zita Sidumo, Paola Germano, Giovanni Guidotti, Carlo Federico Perno

## Abstract

**Background:** Dolutegravir (DTG) is the anchor drug of the first-line agent for HIV treatment globally, including low- and middle-income countries. Although clinical trials report low rates of integrase inhibitor resistance, real-world data from sub-Saharan Africa suggest a different scenario. We aimed to assess the prevalence and patterns of DTG resistance in Mozambique, building on prior preliminary findings with an expanded cohort and extended observation period.

**Methods:** We conducted a retrospective observational study in five DREAM centers in Mozambique. HIV-positive individuals on DTG-based antiretroviral therapy (ART) with confirmed virological failure (HIV RNA >1000 copies/mL) between July 2022 and December 2024 were included. Patients underwent genotypic resistance testing after six months of enhanced adherence support. Resistance mutations were identified via Sanger sequencing and interpreted using the Stanford HIV Drug Resistance Database.

**Results:** Of 28 patients tested, 13 (46.4%) exhibited DTG resistance (12/13 with intermediate or high-level resistance); 92.3% harboured HIV-1 subtype C. Frequently observed mutations included G118R (43.8%), E138K (43.8%), L74M (31.3%), and R263K (25.0%), often in combination. All DTG-resistant individuals who underwent testing for other drug classes (n=9) also showed co-resistance to NRTIs and/or NNRTIs. Notably, high-level resistance emerged also in 6 failing patients having shifted to DTG while virologically suppressed.

**Conclusions:** This study highlights a concerning prevalence of DTG resistance and multiclass failure in Mozambique, underscoring the limitations of current strategies. Sentinel surveillance and expanded access to resistance testing are urgently needed to preserve the efficacy of DTG-based regimens and inform future deployment of long-acting therapies in sub-Saharan Africa.

## Introduction

Despite major global advances in antiretroviral therapy (ART), HIV remains a significant public health challenge, with nearly 40 million people living with the virus worldwide as of 2023 (UNAIDS, 2024). The introduction of integrase strand transfer inhibitors (INSTIs) has transformed HIV treatment by offering potent, well-tolerated regimens with a high genetic barrier to resistance. Among these agents, dolutegravir (DTG) has become the preferred option for first-line ART globally, supported by robust evidence of efficacy, safety, and low rates of treatment-emergent resistance (World Health Organization, 2018). Following the 2018 endorsement by the World Health Organization (WHO), DTG has been rapidly adopted across low- and middle-income countries (LMICs), particularly in sub-Saharan Africa, where the burden of HIV is highest. By 2023, over 90% of LMICs had integrated DTG into their national guidelines, and large-scale roll-out was associated with significant improvements in viral suppression rates (World Health Organization, 2024). In Mozambique—one of the most affected countries, with an estimated 2.4 million people living with HIV—DTG was introduced as part of first-line therapy in 2019 and now represents the backbone of the national ART program (Ministério da Saúde, Direcção Nacional de Saúde Pública, PNC ITS-HIV/SIDA, 2019).

However, growing evidence suggests that the real-world durability of DTG may be challenged in specific populations and contexts. Reports of resistance-associated mutations to DTG are emerging with increasing frequency, particularly in patients with prior ART exposure, poor adherence, or delayed recognition of virological failure (Kamori and Barabona, 2023; Loosli et al., 2023; World Health Organization, 2024). Mutations such as G118R, E138K, and R263K have been identified in multiple settings and are associated with reduced susceptibility to DTG and, in some cases, cross-resistance to other INSTIs (Semengue et al., 2020). These findings are especially concerning in regions with high prevalence of HIV-1 subtype C, where certain resistance pathways may be more accessible to the virus due to subtype-specific polymorphisms. Yet, in countries such as Mozambique, real-world data on resistance patterns remain sparse, and the generalizability of findings from other regions is limited by differences in treatment histories, subtype prevalence, and health system infrastructure.

Although routine viral load monitoring has expanded in recent years, access to genotypic resistance testing remains limited in most LMICs. In Mozambique, surveillance data on DTG resistance are scarce, and little is known about the mutational patterns emerging in patients who fail DTG-based therapy. In the absence of timely resistance surveillance, clinicians are often forced to make empirical decisions regarding regimen switches, which may lead to suboptimal outcomes or missed opportunities to contain emerging resistance. This knowledge gap hampers the timely identification of resistance, the optimization of salvage regimens, and the preservation of DTG’s long-term effectiveness. Elsewhere, we evaluated dolutegravir resistance in a small sample of patients with virological failure in Mozambique, providing one of the first real-world signals of integrase inhibitor resistance in this setting (Doro Altan et al., 2025).

In the present study, we focus exclusively on patients who were receiving DTG-based therapy at the time of resistance testing, extending the observation period, and aim to further investigate the prevalence and patterns of resistance, describe mutational profiles, and explore clinical and programmatic implications in a larger and more comprehensive cohort.

## Methods

### Study Design and Setting

This is a retrospective observational study conducted across five centers of the DREAM program in Mozambique, including two centers in Maputo City, one in Maputo Province, and two in Sofala Province. The DREAM program is a community-based HIV care initiative that integrates clinical follow-up, laboratory monitoring, and social support, and has been active in Mozambique since 2002 (Altan et al., 2016; Ciccacci et al., 2023, 2020; Magnano San Lio et al., 2009).

The current analysis builds on a previously published cohort (Doro Altan et al., 2025), expanding the sample by including all eligible patients who underwent genotypic resistance testing due to confirmed virological failure while on DTG-based ART between July 2022 and December 2024. Compared to the earlier analysis, the present study includes a broader time frame and applies slightly refined inclusion criteria, focusing exclusively on individuals who were receiving DTG at the time of resistance testing, to ensure a more consistent assessment of DTG-associated resistance.

### Study Population

We included HIV-positive individuals on dolutegravir (DTG)-based antiretroviral therapy (ART) who experienced virological failure, defined as HIV RNA >1000 copies/mL, between June 1, 2023, and December 31, 2024. According to DREAM protocols, patients with virological failure were enrolled in a six-month enhanced adherence support program. A repeat viral load test was performed after this period, and those with persistent viremia (>1000 copies/mL) were eligible for genotypic resistance testing. Only individuals on DTG-containing regimens at the time of resistance testing were included in this analysis.

Inclusion criteria were: (i) confirmed HIV-positive status; (ii) being on a DTG-based regimen at the time of resistance testing; (iii) confirmed virological failure (HIV RNA >1000 copies/mL) after at least six months of enhanced adherence support. Exclusion criteria included: (i) switching from DTG to another regimen before resistance testing; (ii) insufficient plasma samples for sequencing; or (iii) incomplete clinical records.

Due to logistical limitations and test availability, only a subset of eligible patients underwent resistance testing.

### Resistance Testing Procedures

Resistance testing followed standardized DREAM protocols. Plasma samples from patients with confirmed virological failure were collected and sent to the Zimpeto DREAM laboratory, where viral RNA was extracted and amplified using an in-house reverse transcription and nested PCR method. Sequencing of the HIV integrase region was performed using the Sanger method at INQABA Biotechnical Industries, South Africa. Sequences were analyzed and interpreted directly at INQABA using the Stanford HIV Drug Resistance Database. DTG resistance was classified according to Stanford penalty scores as follows: susceptible (score <10) (S), low-level resistance (10–14) (LLR), intermediate resistance (15–29) (IR), and high-level resistance (≥30) (HLR). Resistance mutations were identified in the integrase region, and, when available, in the reverse transcriptase and protease regions.

### Data Collection and Variables

Clinical and demographic data were extracted from the DREAM electronic medical records. Variables collected included age, sex, duration on ART, number of prior ART regimens, HIV-1 subtype (when available), and reason for initiating DTG (first-line, switch while suppressed, or switch after failure). Resistance to DTG was classified using Stanford scoring.

### Statistical Analysis

Descriptive statistics were used to summarize baseline characteristics. Categorical variables were presented as frequencies and percentages, and continuous variables as medians with interquartile ranges (IQR).

### Ethics

The study was approved by the National Bioethics Committee for Health of Mozambique (Ref: 430/CNBS/24, July 19, 2024). Given the retrospective nature of the study and the use of routinely collected anonymized data, the requirement for informed consent was waived.

## Results

### Study Population and Baseline Characteristics

In the considered period, a total of 28 consecutive patients on dolutegravir (DTG)-based ART underwent genotypic resistance testing following confirmed virological failure (HIV RNA ≥1000 copies/mL) after six months of enhanced adherence support. Among them, 13 (46.4%) had evidence of resistance to DTG, while 15 (53.6%) remained susceptible.

Table 1 summarizes clinical and demographic characteristics stratified by DTG susceptibility. Patients with DTG resistance were generally older (median age: 38 [IQR: 28-49] vs. 28 years [IQR: 18-40]), had longer treatment histories (median time on ART: 15 [IQR: 9-17] vs. 9 years [IQR: 6-13]), and were more frequently infected with HIV-1 subtype C (92.3% vs. 13.3%) compared to those without resistance. A higher proportion had undergone multiple prior ART regimen changes (61.5% vs. 40.0%). Resistance was observed across all categories of DTG initiation, including patients who started DTG while suppressed (46.2%) or as first-line therapy (7.7%). Among all the 28 patients, 3 received rifampicin-based anti-tuberculosis treatment during DTG treatment, all of them modified DTG dosage according to guidelines; one of them developed DTG resistance.

**Table 1.** Clinical and demographic characteristics of patients undergoing resistance testing, stratified by Dolutegravir (DTG) susceptibility.

| Variable | Total patients tested (n=28) |  | DTG sensible patients (n=15) |  | DTG resistant patients (n=13) |  |
| --- | --- | --- | --- | --- | --- | --- |
| Age (median years, IQR) | 34 | 22-43 | 28 | 18-40 | 38 | 28-49 |
| Sex (n. %) |  |  |  |  |  |  |
| M | 10 | 35,71% | 11 | 73,33% | 9 | 69,23% |
| F | 18 | 64,29% | 4 | 26,67% | 4 | 30,77% |
| Time in ART (median years, IQR) | 11 | 8-15 | 9 | 6-13 | 15 | 9-17 |
| HIV-1 subtype |  |  |  |  |  |  |
| B | 6 | 21,43% | 5 | 33,33% | 1 | 7,69% |
| C | 14 | 50,00% | 2 | 13,33% | 12 | 92,31% |
| unkonwn | 8 | 28,57% | 8 | 53,33% | 0 | 0,00% |
| Reason to DTG therapy |  |  |  |  |  |  |
| In viral failure | 13 | 46,43% | 7 | 46,67% | 6 | 46,15% |
| In viral suppression | 11 | 39,29% | 5 | 33,33% | 6 | 46,15% |
| Start in DTG | 3 | 10,71% | 2 | 13,33% | 1 | 7,69% |
| After abandon | 1 | 3,57% | 1 | 6,67% | 0 | 0,00% |
| Previous regimen changes |  |  |  |  |  |  |
| 0 | 3 | 10,71% | 2 | 13,33% | 1 | 7,69% |
| 1 | 11 | 39,29% | 7 | 46,67% | 4 | 30,77% |
| 2 | 14 | 50,00% | 6 | 40,00% | 8 | 61,54% |

### INSTI Mutational Profiles

Table 2 details the prevalence of specific integrase strand transfer inhibitor (INSTI)-associated mutations among the 13 DTG-resistant individuals. The most frequently observed mutations were G118R and E138K (both in 43.8%), followed by L74M (31.3%) and R263K (25.0%). Other major or accessory mutations, such as Q148R/K and T66A/I, were also detected, often in combination, reflecting complex mutational patterns.

**Table 2.** Frequency of integrase strand transfer inhibitor (INSTI) mutations among patients with DTG resistance.

| Mutation | Occurrence | % of occurrence along the tersted (multiple mutations possible) |
| --- | --- | --- |
| **G118R** | 7 | 43,75% |
| **E138K** | 7 | 43,75% |
| *L74M* | 5 | 31,25% |
| **R263K** | 4 | 25,00% |
| **G140A** | 2 | 12,50% |
| **T66A** | 2 | 12,50% |
| *E157Q* | 2 | 12,50% |
| **E138A** | 1 | 6,25% |
| **Q146P** | 1 | 6,25% |
| **S147G** | 1 | 6,25% |
| **Q148R** | 1 | 6,25% |
| **Q148K** | 1 | 6,25% |
| **T66I** | 1 | 6,25% |
| *S230SR* | 1 | 6,25% |
| *G163R* | 1 | 6,25% |
| *L74I* | 1 | 6,25% |

### DTG Resistance Levels and Co-resistance

Figure 1 illustrates the distribution of DTG resistance levels and associated co-resistance. Based on Stanford interpretation, 1 of the 13 resistant patients had low-level resistance (LLR), 3 had intermediate resistance (IR), and 9 had high-level resistance (HLR). Among the 16 patients tested for resistance to other antiretroviral classes, all were susceptible to protease inhibitors (PI). However, co-resistance was identified in all nine DTG-resistant individuals who underwent testing for other drug classes: eight were resistant to both nucleoside reverse transcriptase inhibitors (NRTIs) and non-nucleoside reverse transcriptase inhibitors (NNRTIs), and one to NRTIs only. The burden of co-resistance increased with the severity of DTG resistance, with multi-drug resistance for NRTI and NNRTI predominantly observed among those with HLR.

**Figure 1.**
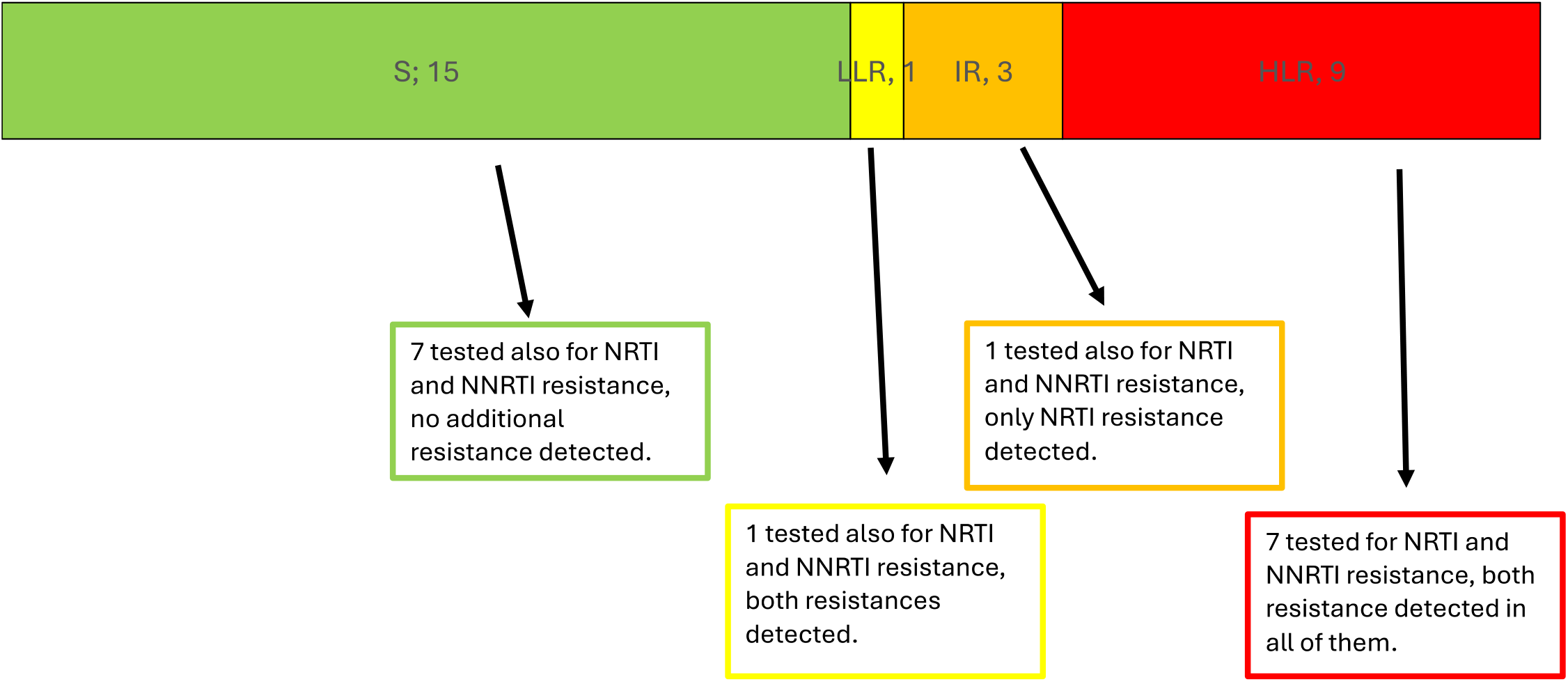
**Distribution of DTG resistance levels according to Stanford classification (S = susceptible, LLR = low-level resistance, IR = intermediate resistance, HLR = high-level resistance). For each resistance category, results of concomitant NRTI and NNRTI resistance testing are shown where available (16 patients underwent testing for additional drug classes).**

### Resistance by Treatment History

Figure 2 presents a Sankey diagram that visualizes the relationship between the reason for initiating DTG and the level of resistance detected in genotypic testing. The diagram shows that HLR emerged across all three groups. Specifically, HLR was identified in individuals who had switched from a failing regimen, but also in patients who started DTG while virologically suppressed and even in those who began DTG as their first-line regimen.

**Figure 2.**
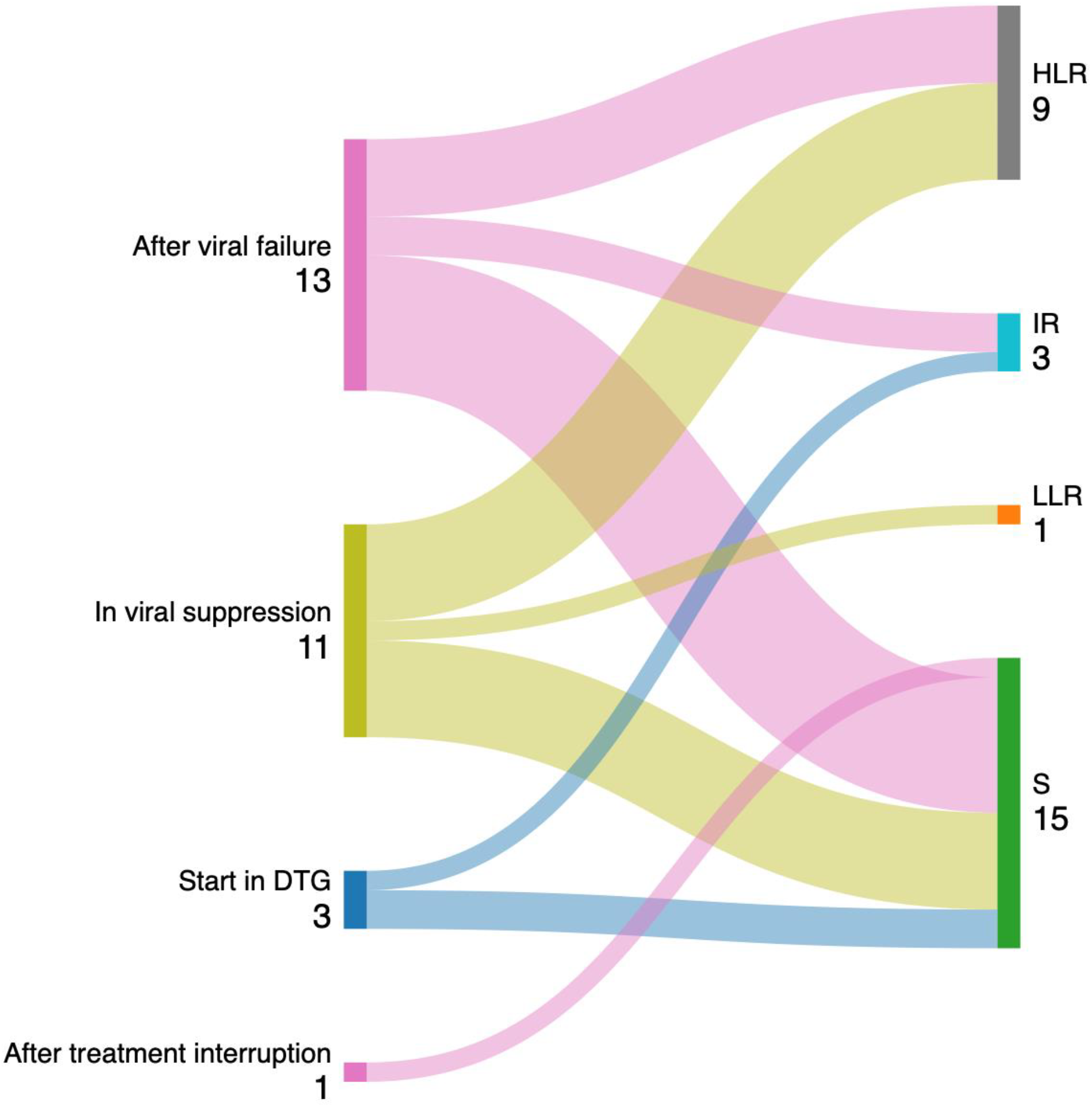
Sankey diagram illustrating the relationship between reason for DTG initiation and level of resistance to DTG.

## Discussion

The results of our study, conducted in five DREAM program centers in Mozambique between 2023 and 2024, reveal a concerning frequency of mutations associated with resistance to DTG among patients with virological failure. Specifically, 46.4% (13 out of 28) of the patients tested for resistance presented integrase-region mutations compatible with reduced DTG susceptibility, with most cases showing intermediate or high-level resistance (12 out of 13). These results align with emerging African literature documenting an increasing incidence of DTG resistance, particularly in real-world settings, contrasting with data from controlled clinical trials.

In large randomized studies such as the NADIA trial (Uganda, Kenya, Zimbabwe) and the DAWNING trial, the rate of INSTI resistance emergence remained low (4% and 1%, respectively), thanks to intensive viral load monitoring and optimized therapeutic strategies (Aboud et al., 2019; Paton et al., 2021). However, these conditions are rarely replicated in routine African healthcare contexts, where virological failures may go undetected for a long period of time. In real-world cohorts similar to ours, such as that reported by Ismael et al. in Mozambique, the prevalence of INSTI mutations among patients with DTG failure reached 15–20% (Ismael et al., 2025); in other cohorts, with different selection criteria, the rates where even higher (Ruano et al., 2025).

The G118R and E138K mutations, each found in 43.8% of our resistant cases, are among the most frequently reported DTG resistance pathways in non-B subtypes, particularly subtype C, which was predominant in our sample. The relevance of G118R has been highlighted by several studies: the meta-analysis by Semengue et al. identified G118R and R263K as the most common INSTI mutations in African failures (Semengue et al., 2020), while the review by Kamori and Barabona emphasized its selection in subtypes A and C, facilitated by its low fitness cost (Kamori and Barabona, 2023). The G118R + R263K combination, also observed in our study, has been associated with high-level resistance and reduced viral fitness (Xiao et al., 2023), and biochemical studies (Quashie et al., 2015) confirmed its preferential selection in subtype C.

Our study also reinforces the strong association between a complex treatment history and resistance risk. DTG-resistant patients had a significantly longer treatment history (median 14.6 years) and more frequent past failures. This is consistent with Loosli et al., who reported similar patterns in a large African-European cohort (Loosli et al., 2023). Furthermore, we observed INSTI mutations even in patients who were DTG-naïve or had initiated the drug while virologically suppressed. This finding are in line with reports from Mahomed et al., Fourie et al., and Mens et al., where G118R emerged under conditions of suboptimal adherence or functionally ineffective regimens (Fourie et al., 2023; Mahomed et al., 2020; Mens et al., 2022).

Another relevant finding is the high rate of co-resistance in our cohort: all patients with INSTI mutations who were tested for other drug classes (9 out of 13) also presented mutations to NRTIs and/or NNRTIs, particularly M184V and K65R, while no resistance to PIs was detected. This pattern mirrors findings from other papers who reported NNRTI and NRTI resistance patients failing TLD (Ismael et al., 2025; Ruano et al., 2025). Our results therefore reinforce the concern that switching to DTG without assessing virological suppression may lead to resistance selection on a background of already compromised backbones.

Lastly, our findings take on special relevance in light of the growing interest in long-acting therapies such as cabotegravir. The G118R mutation, also observed in our study, has been associated with reduced susceptibility to cabotegravir in vitro, raising concerns about cross-resistance (Fokam et al., 2024; Kamori and Barabona, 2023). The editorial by Fokam et al. in Nature Medicine strongly advocates for integrating resistance testing into routine clinical care in low-income countries to avoid silent accumulation of INSTI mutations. Similarly, the review by Tao et al. emphasizes that persistent, unrecognized viremia is a key driver of resistance emergence even in ART-naïve patients, a situation reflected in our cohort (Tao et al., 2023).

### Limitations

This study has several limitations. First, due to the small number of patients included (n=28) and the highly selective nature of the resistance testing, the epidemiological value of our prevalence estimates is limited. Genotypic testing was only performed on patients considered to be at higher risk of harboring resistance, introducing a significant selection bias. This limits the generalizability of our findings to the broader population of patients on DTG in Mozambique.

Second, resistance testing was restricted to the integrase region only, and additional mutations in other regions (e.g., protease or reverse transcriptase) might have gone undetected in patients not fully sequenced. Moreover, Stanford scores were available only for a subset of patients, precluding systematic classification of resistance levels across the full sample.

Finally, data on viral load trajectories, adherence, and treatment history were incomplete for some patients, limiting the capacity to reconstruct the exact pathway leading to the emergence of resistance in each case. Despite these limitations, our study provides important early signals on the possible patterns of DTG resistance in real-world Mozambican settings and highlights the urgent need for expanded molecular surveillance.

### Public Health Implications

The findings of this study have important implications for public health policy and HIV program implementation in sub-Saharan Africa. First, the emergence of DTG resistance— even among patients initiating treatment while virally suppressed—highlights the need to revise current “test-and-switch” strategies. Virological monitoring and resistance testing should be strengthened prior to transitioning patients to DTG-based regimens, especially in settings with a history of limited adherence or prior treatment failures.

Second, our data underscore the urgency of integrating genotypic resistance testing into national HIV guidelines and decentralizing access to such diagnostics. Establishing sentinel surveillance systems in high-prevalence areas could allow for early detection of resistance trends and inform timely changes in treatment protocols.

Finally, the potential for cross-resistance to newer long-acting therapies such as cabotegravir calls for cautious and evidence-based scale-up of such interventions, considering also the extensive resistance to NNRTI typically present in patients having failed first generation NNRTI regimens (World Health Organization, 2024), that might affect the efficacy of rilpivirine (the second drug present in the long-acting regimen). Investments in molecular diagnostic capacity and targeted training of healthcare workers are essential to ensure that therapeutic innovations do not inadvertently accelerate resistance at the population level.

In summary, our data confirm and expand on recent African evidence regarding DTG resistance: G118R, E138K, R263K, and L74M represent emerging mutational patterns in Mozambique, particularly among patients with extensive treatment history, unsuppressed viral load, or prior failures. The silent spread of such mutations, even in clinically stable individuals, poses a tangible risk to the long-term sustainability of DTG-based regimens and highlights the urgent need to implement sentinel genotypic surveillance strategies in public treatment programs.

## Data Availability

All data produced in the present study are available upon reasonable request to the authors

## References

Aboud, M., Kaplan, R., Lombaard, J., Zhang, F., Hidalgo, J.A., Mamedova, E., Losso, M.H., Chetchotisakd, P., Brites, C., Sievers, J., 2019. Dolutegravir versus ritonavir-boosted lopinavir both with dual nucleoside reverse transcriptase inhibitor therapy in adults with HIV-1 infection in whom first-line therapy has failed (DAWNING): an open-label, non-inferiority, phase 3b trial. Lancet Infect. Dis. 19, 253–264.

Altan, A.M.D., Taafo, F., Fopa, F., Buonomo, E., Marazzi, M.C., Nielsen-Saines, K., Orlando, S., Scarcella, P., Ciccacci, F., Mancinelli, S., 2016. An assessment of option B implementation for the prevention of mother to child transmission in Dschang, Cameroon: results from the DREAM (Drug Resource Enhancement against AIDS and Malnutrition) cohort. Pan Afr. Med. J. 23.

Ciccacci, F., Ismael, F., Chume, V., Ruth, L., Mbula, P., Orlando, S., Majid, N.A., Marazzi, M.C., 2023. Enhancing retention in care in HIV-infected adolescents during COVID-19 in Mozambique: results from the DREAM program. Int. J. Adolesc. Med. Health. 10.1515/ijamh-2022-0107

Ciccacci, F., Lucaroni, F., Latagliata, R., Morciano, L., Mondlane, E., Balama, M., Tembo, D., Gondwe, J., Orlando, S., Palombi, L., 2020. Hematologic alterations and early mortality in a cohort of HIV positive African patients. PLoS One 15, e0242068.

Doro Altan, A.M., Majid, N., Orlando, S., Uamusse, E., Rafael, M., Sidumo, Z., Guidotti, G., Ciccacci, F., 2025. Brief communication: virological outcomes and dolutegravir resistance mutations in HIV-infected patients: a multicenter retrospective cohort study in Mozambique. AIDS Res. Ther. 22, 8.

Fokam, J., Inzaule, S., Colizzi, V., Perno, C.-F., Kaseya, J., Ndembi, N., 2024. HIV drug resistance to integrase inhibitors in low-and middle-income countries. Nat. Med. 30, 618–619.

Fourie, N., Rees, K., Mali, D., Mugisa, B., Davies, N., 2023. Dolutegravir resistance in three pregnant and breastfeeding women in South Africa. South. Afr. J. HIV Med. 24.

Ismael, N., Hussein, C., Magul, C., Inguane, H., Couto, A., Nhangave, A., Muteerwa, A., Bonou, M., Ramos, A., Young, P.W., 2025. HIV Drug Resistance Profile in Clients Experiencing Treatment Failure After the Transition to a Dolutegravir-Based First-Line Antiretroviral Treatment Regimen in Mozambique. Pathogens 14, 48.

Kamori, D., Barabona, G., 2023. Dolutegravir resistance in sub-Saharan Africa: should resource-limited settings be concerned for future treatment? Front. Virol. 3, 1253661.

Loosli, T., Hossmann, S., Ingle, S.M., Okhai, H., Kusejko, K., Mouton, J., Bellecave, P., Van Sighem, A., Stecher, M., Monforte, A. d’Arminio, 2023. HIV-1 drug resistance in people on dolutegravir-based antiretroviral therapy: a collaborative cohort analysis. Lancet HIV 10, e733–e741.

Magnano San Lio, M., Mancinelli, S., Palombi, L., Buonomo, E., Altan, A.D., Germano, P., Magid, N.A., Pesaresi, A., Renzi, E., Scarcella, P., Zimba, I., Marazzi, M.C., 2009. The DREAM model’s effectiveness in health promotion of AIDS patients in Africa. Health Promot Int 24, 6–15. 10.1093/heapro/dan043

Mahomed, K., Wallis, C.L., Dunn, L., Maharaj, S., Maartens, G., Meintjes, G., 2020. Case report: Emergence of dolutegravir resistance in a patient on second-line antiretroviral therapy. South. Afr. J. HIV Med. 21.

Mens, H., Fjordside, L., Fonager, J., Gerstoft, J., 2022. Emergence of the G118R pan-integrase resistance mutation as a result of low compliance to a dolutegravir-based cART. Infect. Dis. Rep. 14, 501–504.

Ministério da Saúde, Direcção Nacional de Saúde Pública, PNC ITS-HIV/SIDA, 2019. Normas Clínicas Actualizadas para o seguimento do paciente HIV positivo.

Paton, N.I., Musaazi, J., Kityo, C., Walimbwa, S., Hoppe, A., Balyegisawa, A., Kaimal, A., Mirembe, G., Tukamushabe, P., Ategeka, G., 2021. Dolutegravir or darunavir in combination with zidovudine or tenofovir to treat HIV. N. Engl. J. Med. 385, 330–341.

Quashie, P.K., Oliviera, M., Veres, T., Osman, N., Han, Y.-S., Hassounah, S., Lie, Y., Huang, W., Mesplède, T., Wainberg, M.A., 2015. Differential effects of the G118R, H51Y, and E138K resistance substitutions in different subtypes of HIV integrase. J. Virol. 89, 3163–3175.

Ruano, M., Flores, A., Couto, A., Gaspar, I., Yerly, S., Zamudio, A.G.G., Bene, R., Maiela, A., Macuacua, H., Lane, J., 2025. Dolutegravir Resistance in Mozambique: Insights from a Programmatic HIV Resistance Testing Intervention in a Highly ART Experienced Cohort.

Semengue, E.N.J., Santoro, M.M., Ndze, V.N., Dambaya, B., Takou, D., Teto, G., Nka, A.D., Fabeni, L., Wiyeh, A.B., Ceccherini-Silberstein, F., Colizzi, V., Perno, C.F., Fokam, J., 2020. HIV-1 Integrase Resistance Associated Mutations and the Use of Dolutegravir in Sub-Saharan Africa: A Systematic Review and Meta-Analysis Protocol. Syst. Rev. 10.1186/s13643-020-01356-z

Tao, K., Rhee, S.-Y., Chu, C., Avalos, A., Ahluwalia, A.K., Gupta, R.K., Jordan, M.R., Shafer, R.W., 2023. Treatment emergent dolutegravir resistance mutations in individuals naive to HIV-1 integrase inhibitors: a rapid scoping review. Viruses 15, 1932.

UNAIDS, 2024. Global HIV & AIDS statistics — Fact sheet [WWW Document]. URL https://www.unaids.org/en/resources/fact-sheet (xaccessed 10.10.24).

World Health Organization, 2024. HIV drug resistance: brief report 2024.

World Health Organization, 2018. Updated recommendations on first-line and second-line antiretroviral regimens and post-exposure prophylaxis and recommendations on early infant diagnosis of HIV: interim guidelines. Supplement to the 2016 consolidated guidelines on the use of antiretroviral drugs for treating and preventing HIV infection.

Xiao, M.A., Cleyle, J., Yoo, S., Forrest, M., Krullaars, Z., Pham, H.T., Mesplède, T., 2023. The G118R plus R263K combination of integrase mutations associated with dolutegravir-based treatment failure reduces HIV-1 replicative capacity and integration. Antimicrob. Agents Chemother. 67, e01386–22.

